# Integrative transcriptomic analysis identifies long noncoding RNA dysregulation and circadian disruption in reward and executive circuits of opioid use disorder

**DOI:** 10.64898/2026.02.14.26346327

**Authors:** Zixiu Li, Chen Fu, Peng Zhou, Ryan W Logan, Chan Zhou

**Affiliations:** Population and Quantitative Health Sciences, University of Massachusetts Chan Medical School, Worcester, MA, USA 01655; Department of Psychiatry and Behavioral Sciences, University of Massachusetts Chan Medical School, Worcester, MA, USA 01655; Department of Neurobiology, University of Massachusetts Chan Medical School, Worcester, MA, USA 01655; Systems, Computational, and Quantitative Biology, Morningside Graduate School of Biomedical Sciences, UMass Chan Medical School, Worcester, MA, USA 01655; The RNA Therapeutics Institute, University of Massachusetts Chan Medical School, Worcester, MA, USA 01655

**Author notes:** These authors contributed equally to this work (Zixiu Li, Chen Fu). Correspondence: RWL and CZ.

## Abstract

Opioid use disorder (OUD) is characterized by compulsive drug seeking and impaired executive control arising from maladaptive plasticity within cortico–striatal circuits. While transcriptomic studies have identified coding gene alterations in the nucleus accumbens (NAc) and dorsolateral prefrontal cortex (DLPFC), the contribution of the noncoding genome remains poorly defined. Here, we performed integrative transcriptomic analysis of postmortem human NAc and DLPFC to systematically identify and characterize long noncoding RNAs (lncRNAs) in OUD. We identified 36,225 lncRNA loci expressed across reward and executive regions, approximately half of which were previously unannotated. OUD was associated with widespread lncRNA dysregulation in NAc and DLPFC, with lncRNA-centered co-expression modules enriched for neuroimmune signaling, phosphorylation-dependent synaptic pathways, and intracellular receptor cascades. Notably, OUD disrupted circadian rhythmicity of lncRNAs to a degree comparable to or exceeding mRNAs, implicating temporal reorganization of noncoding networks in addiction pathology. Integration with single-nucleus transcriptomic data revealed pronounced neuronal and glial cell type specificity among OUD-associated lncRNAs. Together, these findings demonstrate that lncRNAs represent a critical regulatory layer in reward and executive circuits and suggest that spatial, temporal, and cellular remodeling of the noncoding transcriptome contributes to circuit dysfunction in OUD.

## INTRODUCTION

Opioid use disorder (OUD) is a chronic, relapsing brain disease marked by compulsive drug seeking, impaired executive control, and a high propensity for relapse. These core behavioral features arise from maladaptive plasticity within cortico–striatal circuitry, particularly the nucleus accumbens (NAc), which integrates reward processing and motivational salience, and the dorsolateral prefrontal cortex (DLPFC), which mediates executive function, decision-making, and behavioral inhibition^1^. Repeated opioid exposure reshapes these interconnected regions, leading to persistent alterations in synaptic signaling and circuit dynamics that underlie compulsive use and loss of control.

Large-scale transcriptomic and proteomic profiling of postmortem human brain has begun to elucidate the molecular architecture of these adaptations. Prior studies, including our own, of the NAc and DLPFC in individuals with OUD have consistently identified dysregulation of neuroinflammatory pathways, extracellular matrix remodeling, phosphorylation-dependent synaptic signaling, and circadian rhythm– associated programs^2–6^. However, these investigations have focused predominantly on protein-coding transcripts and proteins, leaving the noncoding transcriptome—particularly long noncoding RNAs (lncRNAs)—largely unexplored. Consequently, the contribution of the noncoding genome to circuit-level dysfunction in OUD remains insufficiently defined.

Long noncoding RNAs (lncRNAs) represent a vast and highly tissue-specific layer of the human transcriptome^7^. Constituting the majority of transcribed genomic loci, lncRNAs regulate gene expression through diverse mechanisms, including chromatin modification, transcriptional modulation, RNA–protein scaffolding, and post-transcriptional regulation. They often exhibit tissue- and disease-specific expression patterns^8^. Accumulating evidence demonstrated that lncRNAs participate in multiple foundamental biological processes and disease progression^9–18^, The nervous system expresses an unusually rich repertoire of lncRNAs, many of which exhibit pronounced brain-region and cell-type specificity and dynamic regulation during neuronal activity and synaptic plasticity^19–21^. Increasing evidence demonstrates that lncRNAs may participate in neuronal differentiation, synaptic function, and circuit regulation^19–21^. Despite these advances, the roles of lncRNAs in opioid use disorder remain largely unexplored.

Several factors have limited the research progress of lncRNAs in human brain disorders. First, current genome annotations incompletely capture brain-expressed lncRNAs. Our prior work indicates that up to 50–60% of lncRNAs detected in individual human samples are not represented in existing annotations^22^. Second, most addiction transcriptomics studies have relied on annotation-dependent pipelines, potentially overlooking novel lncRNAs. Third, the rhythmic and circuit-level dynamics of lncRNAs in OUD have not been systematically examined. Circadian dysregulation has emerged as a key feature of OUD, yet whether lncRNAs participate in rhythmic reorganization of reward and executive circuits remains unknown. Finally, the cell type specificity of OUD-associated lncRNAs has not been comprehensively defined.

Thus, a critical gap remains in understanding how lncRNAs contributes to molecular adaptations in OUD, particularly within reward (NAc) and executive control (DLPFC) circuits.

To address this gap, we performed a comprehensive identification and characterization of lncRNAs in postmortem human NAc and DLPFC by integrative bioinformatics data analysis to define the landscape and potential roles of lncRNAs in OUD. We identified thousands of previously unannotated brain-expressed lncRNAs, revealed OUD-associated lncRNA dysregulation in both regions, demonstrated disruption of lncRNA circadian rhythmicity, and uncovered pronounced neuronal and glial cell type specificity in striatal circuits. Our findings position lncRNAs as integral components of molecular networks underlying reward dysregulation and executive dysfunction in OUD.

## RESULTS

### Many lncRNAs are widely expressed with diverse genomic origins in nucleus accumbens (NAc) and dorsolateral prefrontal cortex (DLPFC)

To explore lncRNAs in OUD, we first systematically identified the landscape of lncRNAs expressed in NAc and DLPFC, two key brain regions critical for reward processing and executively control over drug-seeking behavior^23,24^. Using our previously developed lncRNA detection method, *Flnc*^22^, we analyzed polyA-selected RNA-seq data from 223 post-mortem NAc samples and 30 DLPFC samples. Across individual samples, we detected thousands of lncRNAs **(Figure 1A-1B**), with ∼ 40-70% classified as novel in both brain regions (**Figure 1A-1B**). The sample-to-sample variation in detected lncRNA numbers largely tracked sequencing depth, due to the generally lower expression of lncRNAs compared with mRNAs in the tissue level.

**Figure 1.**
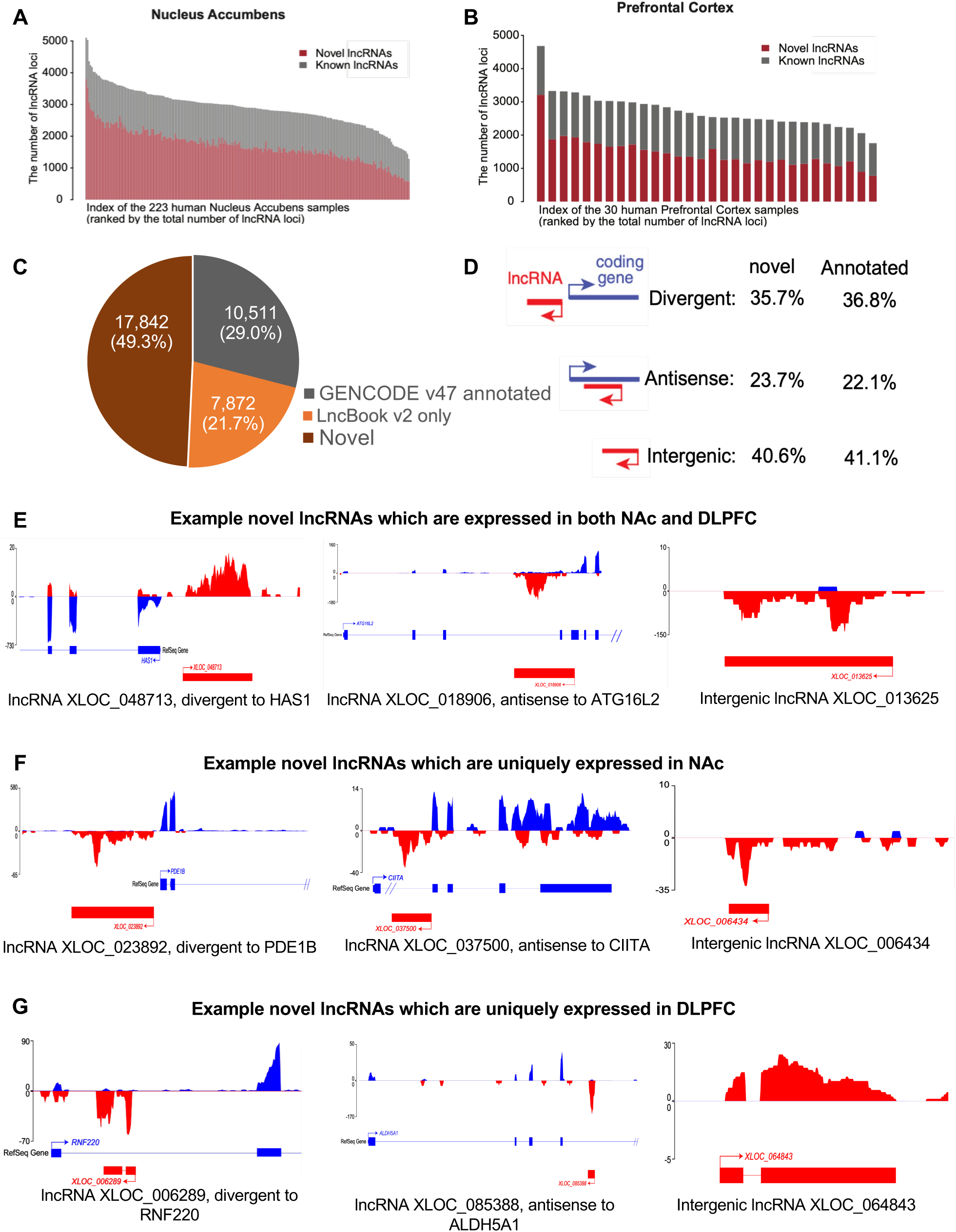
Full landscape of novel and annotated lncRNAs in human reward (NAc) and executive control circuit regions (DLPFC). Comprehensive profiling of long noncoding RNAs (lncRNAs) in post-mortem nucleus accumbens (NAc; n=223) and dorsolateral prefrontal cortex (DLPFC; n=30) reveals a large repertoire of previously unannotated transcripts in brain regions central to reward processing and executive control. (A) Number of novel (red) and annotated (grey) lncRNAs detected in each NAc sample, illustrating substantial inter-individual variability in lncRNA repertoire within the reward circuitry. (B) Number of novel (red) and annotated (grey) lncRNAs detected in each DLPFC sample, demonstrating robust lncRNA expression in cortical regulatory regions. (C) Overall distribution of novel and annotated lncRNAs identified from polyA RNA-seq data of post-mortem NAc and DLPFC samples, highlighting the substantial fraction of previously uncharacterized brain-expressed lncRNAs. (D) Genomic origins of novel and annotated lncRNA loci. Divergent lncRNAs are defined as transcripts with a transcription start site (TSS) located within ±2 kb of the TSS of a protein-coding gene on the opposite strand. Antisense lncRNAs overlap a protein-coding gene by at least one base pair on the opposite strand. All remaining lncRNA transcripts were classified as intergenic. lncRNAs are shown in red and protein-coding genes in grey. The abundance of each category is indicated on the right. (E) Representative examples of novel lncRNAs expressed in both NAc and DLPFC brain regions. XLOC_048713 is divergent to HAS1 (left), XLOC_018906 is antisense to ATG16L2 (middle), and XLOC_013625 is an intergenic lncRNA (right). RNA-seq coverage is shown in red for lncRNAs and blue for protein-coding genes. Gene models are displayed below (exons as boxes, introns as lines, arrows indicating transcription start site and direction).Double slashes (“//”) denote omitted genomic regions containing multiple exons. (F) Representative examples of novel lncRNAs uniquely expressed in NAc. XLOC_023892 is divergent to PDE1B (left), XLOC_037500 is antisense to CIITA (middle), and XLOC_006434 is an intergenic lncRNA (right). RNA-seq read coverage tracks and gene models are shown as in panel E. (G) Representative examples of novel lncRNAs uniquely expressed in DLPFC. XLOC_006289 is divergent to RNF220 (left), XLOC_ 085388 is antisense to ALDH5A1 (middle), and XLOC_064843 is an intergenic lncRNA (right). RNA-seq read coverage tracks and gene models are shown as in panel E.

In total, we identified 36,225 lncRNA loci with expression in at least one sample across these two brain regions. Approximately half of these lncRNA loci were not previously reported, whereas 29% were curated in GENCODE^25,26^ and 21.7% were predicted loci collected in the LncBook database^27^ (**Figure 1C**). Notably, the fraction of novel lncRNA transcripts (isoforms) exceeded the fraction of novel loci, reflecting substantial novel lncRNA-isoform discovery from annotated/known loci in NAc and DLPFC.

We next examined the genomic origins of these lncRNAs. The overall pattern was similar between novel and annotated lncRNAs: > 35% of lncRNAs were divergent from protein-coding genes, ∼ 23% were antisense to coding genes, and > 40% originated from intergenic regions (**Figure 1D**). The relatively high proportion of intergenic lncRNAs in these brain regions appears greater than that observed in several non-brain tissues^28,29^. This may suggest the possibility that human brain harbors a particularly rich repertoire of brain-region- and cell type--specific lncRNAs.

To illustrate these three genomic categories and the confidence of our novel lncRNAs, we visualized RNA-seq read coverage support and exon structures for representative novel lncRNAs from each category using genome browser tracks. We highlight examples expressed in both NAc and DLPFC (**Figure 1E**), NAc-specific lncRNAs (**Figure 1F**), and DLPFC-specific lncRNAs (**Figure 1G**). Together, these results establish a large catalog of annotated and novel lncRNAs in OUD-relevant human brain regions and demonstrate pronounced regional specificity at the transcript and loci levels.

### Opioid use disorder dysregulated lncRNA expression in the nucleus accumbens

To reveal potential lncRNA contributions to OUD in reward circuitry, we performed differential expression analysis across 36,225 detected lncRNAs using post-mortem NAc samples from 20 OUD subjects with OUD and 20 unaffected control subjects (See Methods). This analysis found 375 lncRNAs with OUD-associated expression changes, of which ∼45% were novel (**Figure 2A**). These differentially expressed (DE) lncRNAs include 181 OUD-induced and 194 OUD-repressed lncRNAs (**Figure 2B**).

**Figure 2.**
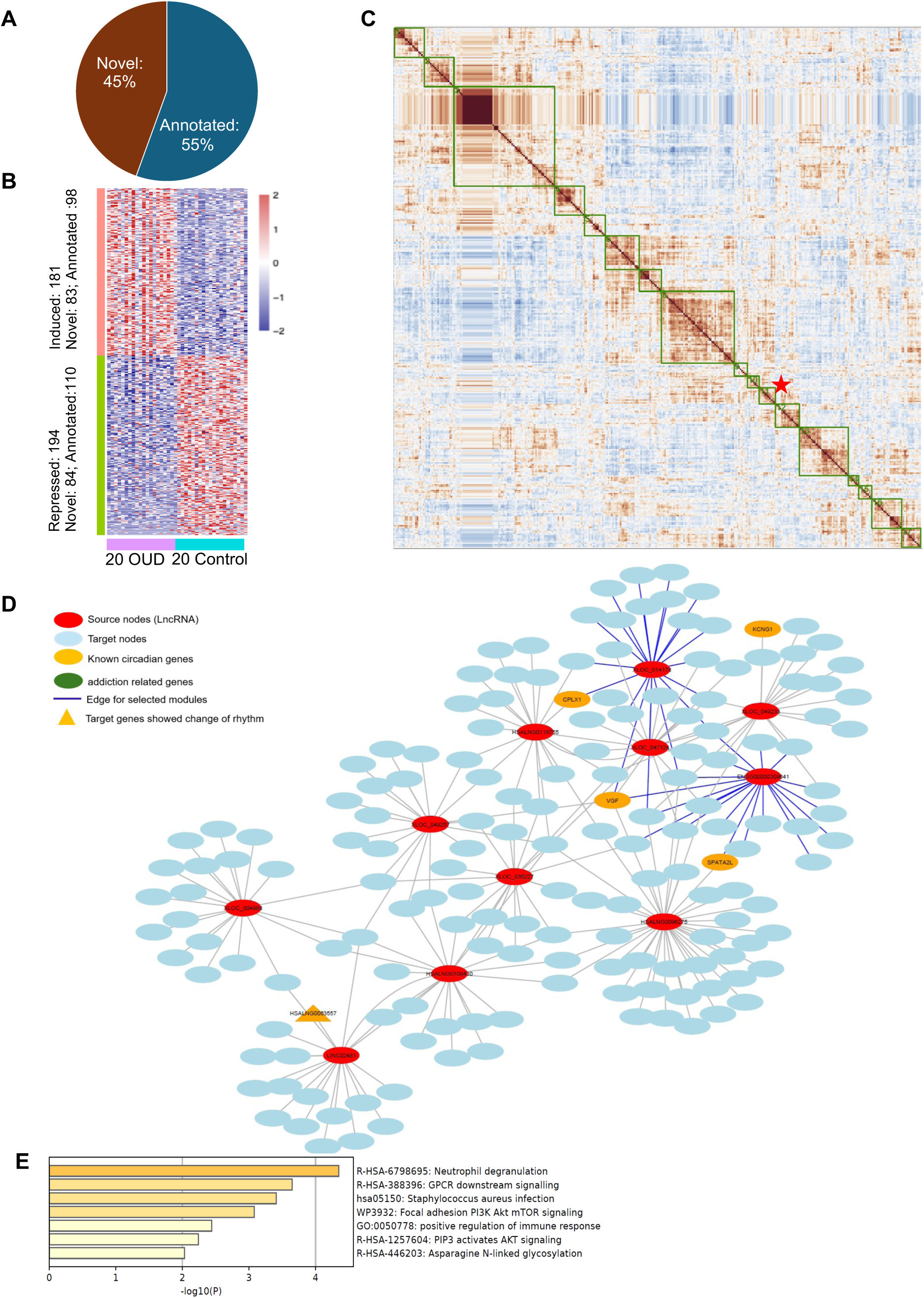
Human lncRNAs are dysregulated in the nucleus accumbens of individuals with opioid use disorder. (A) Proportion of differentially expressed lncRNAs in NAc (20 OUD vs. 20 controls), stratified as novel or annotated. (B) Heatmap of differentially expressed lncRNAs across 20 OUD and 20 control samples. Rows represent lncRNAs and columns represent individuals. “Induced” and “Repressed” indicate higher or lower expression in OUD relative to controls, respectively. (C) Correlation matrix of co-expressed networks seeded with differentially expressed (DE) lncRNAs. Networks with high similarity were grouped together into modules (outlined in green). For each module, an addition-enrichment score was quantified as the proportion of addiction- or circadian-related genes among its protein-coding genes in the module. The module with the strongest addition-related enrichment, suggesting lncRNA-centered regulatory programs linked to reward or circadian dysregulation in OUD, was marked with a red asterisk. (D) Co-expression networks between DE lncRNAs and coding genes for representative module 13. The co-expression among coding genes were not shown here. Red nodes indicate DE lncRNAs; blue nodes indicate co-expressed protein-coding genes. Yellow nodes denote known circadian-related genes, and green nodes denote addiction-related genes, highlighting the integration of lncRNAs within biologically relevant regulatory circuits. (E) Pathway enrichment analysis of protein-coding genes co-expressed with DE lncRNAs in module 13, revealing neurological immune and signaling pathways implicated in OUD-related neurobiological adaptations.

To infer putative functional roles of these OUD-regulated lncRNAs, we further performed lncRNA-centered co-expression network analysis (see Methods). Briefly, each DE lncRNA was treated as a seed/hub to define a local co-expression neighborhood of protein-coding genes, and these local co-expression networks were grouped into a higher-order modules based on functional similarity among their coding-gene components (See Methods). This analysis yielded 17 co-expression modules for OUD-regulated lncRNAs in NAc (**Figure 2C**). We ranked modules for relevance to opioid addiction by the proportion of addiction- and circadian-related genes among their protein-coding genes. Module 11 showed the strongest addition-related enrichment (**Figure 2C**, red asterisk), suggesting lncRNA-centered regulatory programs linked to core addiction biology in NAc.

Network visualization of Module 11 (**Figure 2D**) highlights co-expression relationships between DE lncRNAs (red) and circadian-related genes (coding genes in orange oval; lncRNAs in orange triangles). Pathway enrichment analysis for Module 11 implicated multiple biological processes, including positive regulation of immune response and neutrophil degranulation. It suggested that a subset of OUD-regulated lncRNAs may participate in neuroimmune signaling programs within NAc. In parallel, Module 11’s enrichment in genes related to focal adhesion PI3K AKT mTOR signaling, GPCR downstream signaling, and PIP3 activates AKT signaling suggested these lncRNAs in Module 11 may also be involved in synapse signaling and plasticity implicated in opioid addiction.

### Opioid use disorder dysregulated lncRNA expressions in the dorsolateral prefrontal cortex

Because the DLPFC is a critical executive control region implicated in decision-making, impulsivity, top-down control of drug seeking, we performed differential expression analysis in DLPFC using post-mortem samples from OUD subjects and controls (see Methods). We identified 102 lncRNAs dysregulated in DLPFC, including 37% novel lncRNAs (**Figure 3A**). These comprised 56 OUD-induced and 46 OUD-repressed lncRNAs (**Figure 3B**).

**Figure 3.**
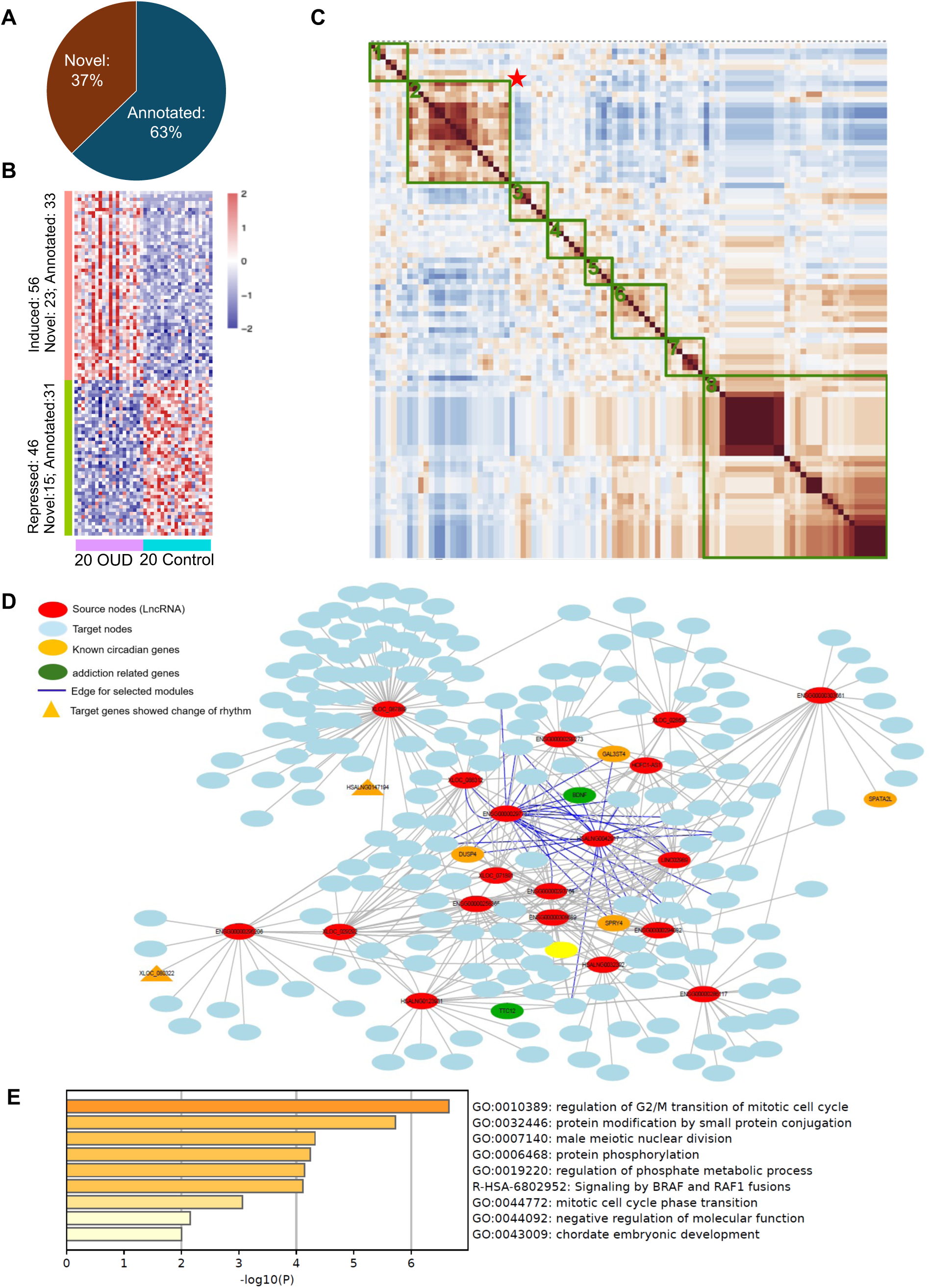
Human lncRNAs are dysregulated in the dorsolateral prefrontal cortex (DLPFC) of individuals with opioid use disorder (OUD). (A) Distribution of differentially expressed lncRNAs in DLPFC (20 OUD vs. 20 controls), stratified as novel and annotated transcripts. (B) Heatmap of differentially expressed lncRNAs across DLPFC samples. Rows represent lncRNAs and columns represent individuals. “Induced” and “Repressed” denote higher or lower expression in OUD relative to controls, respectively. (C) Correlation matrix of co-expression networks seeded with differentially expressed (DE) lncRNAs. Highly similar networks were grouped into modules (outlined in green). Enrichment was quantified as the proportion of addiction- or circadian-related genes among protein-coding members of each module. Modules with the strongest addiction-related enrichment, indicating lncRNA-centered regulatory programs potentially contributing to executive dysfunction and reward circuitry alterations in OUD, are marked with a red asterisk. (D) Co-expression network of a representative addiction-enriched module. Red nodes denote DE lncRNAs; blue nodes represent co-expressed protein-coding genes. Yellow nodes indicate circadian genes, and green nodes indicate addiction-related genes, highlighting integration of lncRNAs within disease-relevant regulatory circuits. (E) Pathway enrichment analysis of protein-coding genes co-expressed with DE lncRNAs in the selected module, revealing pathways related to cell-cycle regulation and signaling processes implicated in cortical plasticity and OUD-associated neuroadaptation.

Using the same co-expression module framework applied in NAc, we identified eight lncRNAs-centered co-expression modules for OUD-regulated lncRNAs in DLPFC (**Figure 3C**). Among these eight modules, Module 2 showed the strongest addition-related enrichment. In the Module 2 network (**Figure 3D),** DE lncRNAs co-expressed with both addiction-related genes (green) and circadian-related genes (orange), reinforcing cross-talk between addiction pathways and rhythmic biology in executive control circuitry^2–4^.

Pathway analysis of DLPFC Module 2 highlighted protein phosphorylation and regulation of phosphate metabolic processes, aligning with phosphorylation-centered signaling themes observed in NAc Module 11 (including PI3K–AKT–mTOR signaling and IP3→AKT activation. Given that PI3K–AKT–mTOR signaling is fundamentally driven by phosphorylation cascades, this cross-region concordance supports phosphorylation-dependent signaling as a recurring molecular feature associated with OUD.

In addition to shared signaling pathways, DLPFC Module showed significant enrichment for mitotic cell cycle transition and G2/M regulation, which were not observed among the significant pathways in NAc Module 11. Because neurons are largely post-mitotic, these signals likely reflect contributions from non-neuronal cell populations (e.g., glia). This finding raises the possibility that glia-associated proliferative or cell-cycle dynamics may be preferentially engaged in the DLPFC, but not in the NAc, in opioid addiction.

### Opioid use disorder disrupts lncRNA circadian rhythmicity in reward and prefrontal circuits

Prior studies, including our work^2,4^, suggested that molecular rhythm disruptions in brain regions, including DLPFC and NAc, are key factors in opioid addiction. Consistent with this, we observed that subsets of OUD-regulated DE lncRNAs co-expressed with circadian genes in both NAc (**Figure 2D**) and DLPFC (**Figure 3D**), suggesting that circadian rhythmicity of lncRNAs might be disrupted in opioid addition. Therefore, we performed differential rhythmicity (DR) analysis for lncRNAs in both brain regions.

We identified 83 lncRNAs in NAc and 109 lncRNAs in DLPFC with significant alterations in rhythmicity between OUD and unaffected controls. Notably, these DR lncRNAs outnumbered DR mRNAs (56 in NAc and 73 in DLPFC) (**Figure 4A**). DLPFC exhibited more rhythmicity-altered lncRNAs and mRNAs than NAc, suggesting comparatively stronger rhythmic disruption in executive circuitry (Figure 4A). Together, these results indicate that OUD-associated circadian disruption extends robustly to the lncRNA transcriptome, at least comparable to protein-coding genes’ rhythm disruption.

**Figure 4.**
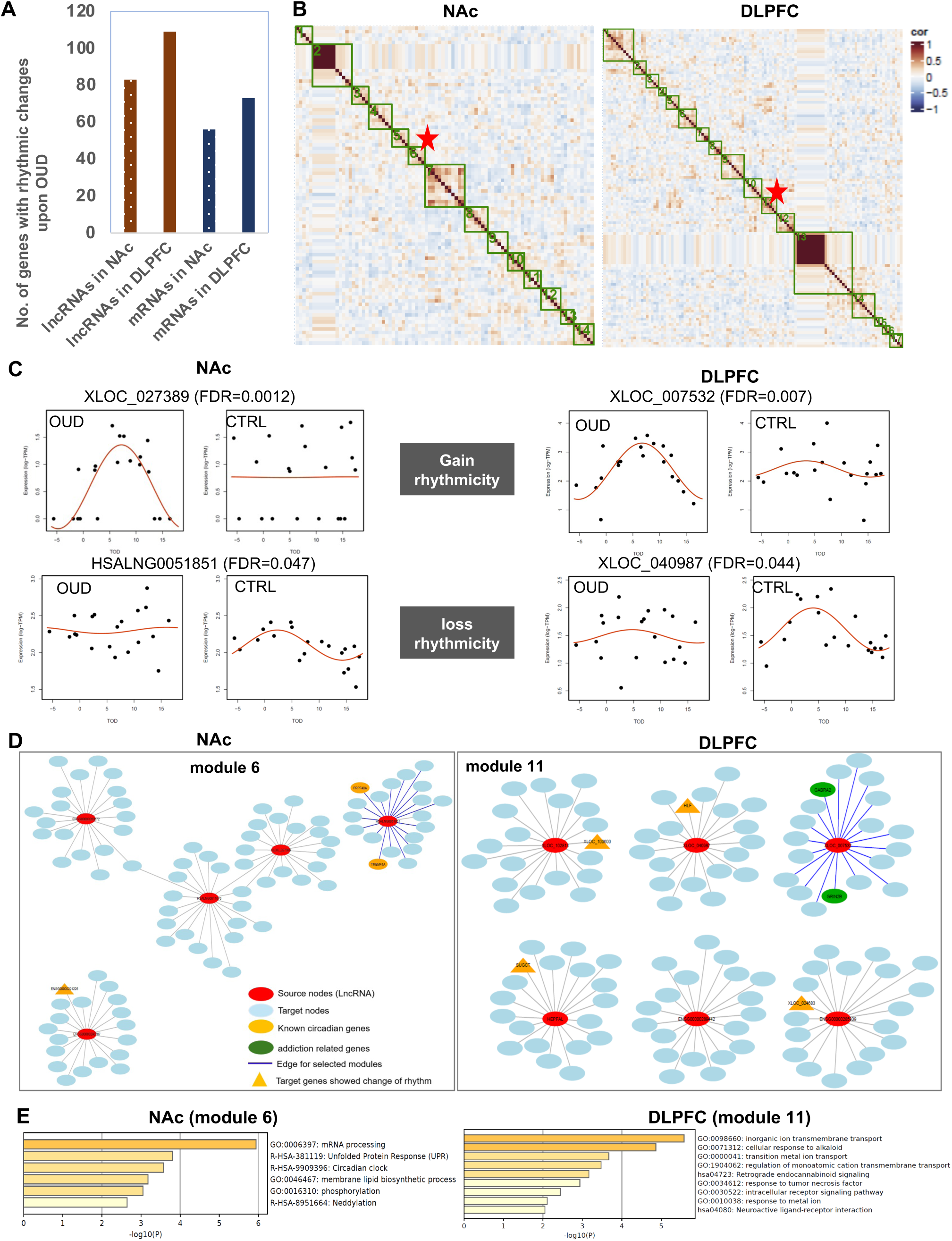
Opioid use disorder disrupts lncRNA circadian rhythmicity in reward and prefrontal circuits. (A) Number of lncRNAs and mRNAs exhibiting significant changes in circadian rhythmicity in the nucleus accumbens (NAc) and dorsolateral prefrontal cortex (DLPFC) of individuals with OUD relative to controls. (B) Correlation matrices of co-expression networks seeded with lncRNAs showing altered rhythmicity in NAc and DLPFC. Highly similar networks were grouped into modules (outlined in green). Modules enriched for addiction- or circadian-related genes are marked with a red asterisk. (C) Representative examples of lncRNAs displaying gain or loss of rhythmicity in OUD compared with controls in NAc and DLPFC. FDR values are indicated. Curves represent fitted rhythmic patterns. (D) Co-expression network visualization of representative rhythmicity-associated modules (NAc module 6; DLPFC module 11). Red nodes denote lncRNAs; blue nodes represent co-expressed protein-coding genes; yellow nodes indicate circadian genes; green nodes denote addiction-related genes; triangles mark genes exhibiting altered rhythmicity. Networks highlight region-specific lncRNA-centered circuits: in NAc module 6, rhythmicity-altered lncRNAs co-express primarily with circadian genes, whereas in DLPFC module 11 they co-express with circadian and addiction-related genes, linking rhythmic disruption to addiction-relevant programs. (E) Pathway enrichment analysis of protein-coding genes within the selected rhythmic modules in NAc and DLPFC, suggesting region-specific consequences of the rhythmic-altered lncRNAs in OUD.

While OUD disrupted lncRNA circadian rhythmicity in NAc and DLPFC, some lncRNAs significantly gained rhythmicity in opioid addiction, such as XLOC_027389 in NAc and XLOC_007532 in DLPFC, whereas some lncRNAs showed lost rhythmicity in opioid addiction, such as HSALNG0051851 in NAc and XLOC_040987 in DLPFC (**Figure 4C**).

To infer potential functional roles of DR lncRNAs, we constructed co-expression modules seeded by rhythmicity-altered lncRNAs, analogous to the DE-lncRNA co-expression module analysis. Several modules were prioritized (**Figure 4B**). In NAc, Module 6 showed co-expression between HSALN051851, a DR lncRNA which lost rhythmicity in OUD, and two established circadian-related genes, PRPF40A and TMEM41A. This observation suggested that its altered oscillatory dynamics might be linked to dysregulation of co-expressed circadian genes within this module. Consistently, pathway analysis of NAc Module 6 identified *Circadian clock* among top enriched terms (**Figure 4E, left**), supporting functional convergence of this module on circadian regulation. Notably, phosphorylation-related pathways were also enriched, echoing network patterns observed for DE lncRNAs and suggesting that phosphorylation-dependent signaling may intersect lncRNA-involved circadian dysregulation in OUD.

In DLPFC, XLOC_007532 gained rhythmicity in opioid addiction and was strongly co-expressed with two well-established addiction-related genes, GRIN2B and GABRA2 (**Figure 4D, right**) within Module 11. This pattern implied that altered rhythmic regulation of this lncRNA might be associated with oscillatory changes in excitatory and inhibitory neurotransmission. Within DLPFC Module 11, four of five additional DR lncRNAs were co-expressed with mRNAs (HLF, SUGCT) or lncRNAs (*XLOC_10060*, *XLOC_026483*) that also exhibited rhythmic alterations, indicating coordinated oscillatory reprogramming within this network. Consistent with this interpretation, pathway enrichment of DLPFC Module 11 (Figure 4E, right) revealed significant overrepresentation of processes related to inorganic ion transmembrane transport, transition metal ion transport, and regulation of monoatomic cation transmembrane transport, along with retrograde endocannabinoid signaling, neuroactive ligand–receptor interaction, and intracellular receptor signaling pathways. Enrichment for response to tumor necrosis factor and cellular response to alkaloid further suggests integration of inflammatory and opioid-related signaling within this rhythmic module.

Collectively, these findings suggest OUD may reshape the circadian coordination of lncRNA–mRNA networks in DLPFC, converging on ion transport, receptor signaling, and neuro-modulatory pathways central to executive control circuitry.

### OUD-associated lncRNAs exhibit cell type–specific expression patterns in the striatum

Because lncRNAs are often highly cell type-specific and brain regions are cellularly heterogenous, we next investigated the cell type specificity of OUD-dysregulated lncRNAs. Due to limited availability of OUD-matched single-cell datasets from NAc and DLPFC, we re-analyzed our previously generated single-nucleus RNA-seq (snRNA-seq) data from the dorsal striatum^30^, leveraging the shared cellular composition between dorsal striatum and NAc (ventral striatum) as a proxy tissue for cell type–resolved inference.

Using the same cell type annotation as our prior work for this snRNA-seq data^30^, the striatum cells are mainly grouped into neurons (D1-MSNs, D2-MSNs and interneurons), glial cells (astrocytes, oligodendrocyte lineage cells), vascular cells (mural and endothelial), and microglia (**Figure 5A**). In total, 64 OUD-dysregulated lncRNAs exhibited cell type–specific expression patterns, including 44 OUD-induced lncRNAs and 20 OUD-repressed lncRNAs (**Figure 5B)**. Among the 44 induced lncRNAs with cell type specificity, 29 (66%) were enriched in neurons, 6 were enriched in both neurons and glial cells but minimal expressed in microglia, and 4 were specifically enriched in microglia **(Figure 5B, left)**. Among 20 repressed lncRNAs, 9 were enriched in microglia, 5 in astrocytes, 4 in neurons, and 1 in D1-MSNs **(Figure 5B, right)**. These cell-type specific patterns were supported by cell coverages heatmap across major striatal cell types **(Figure 5C),** which were largely consistent between OUD and non-OUD subjects.

**Figure 5.**
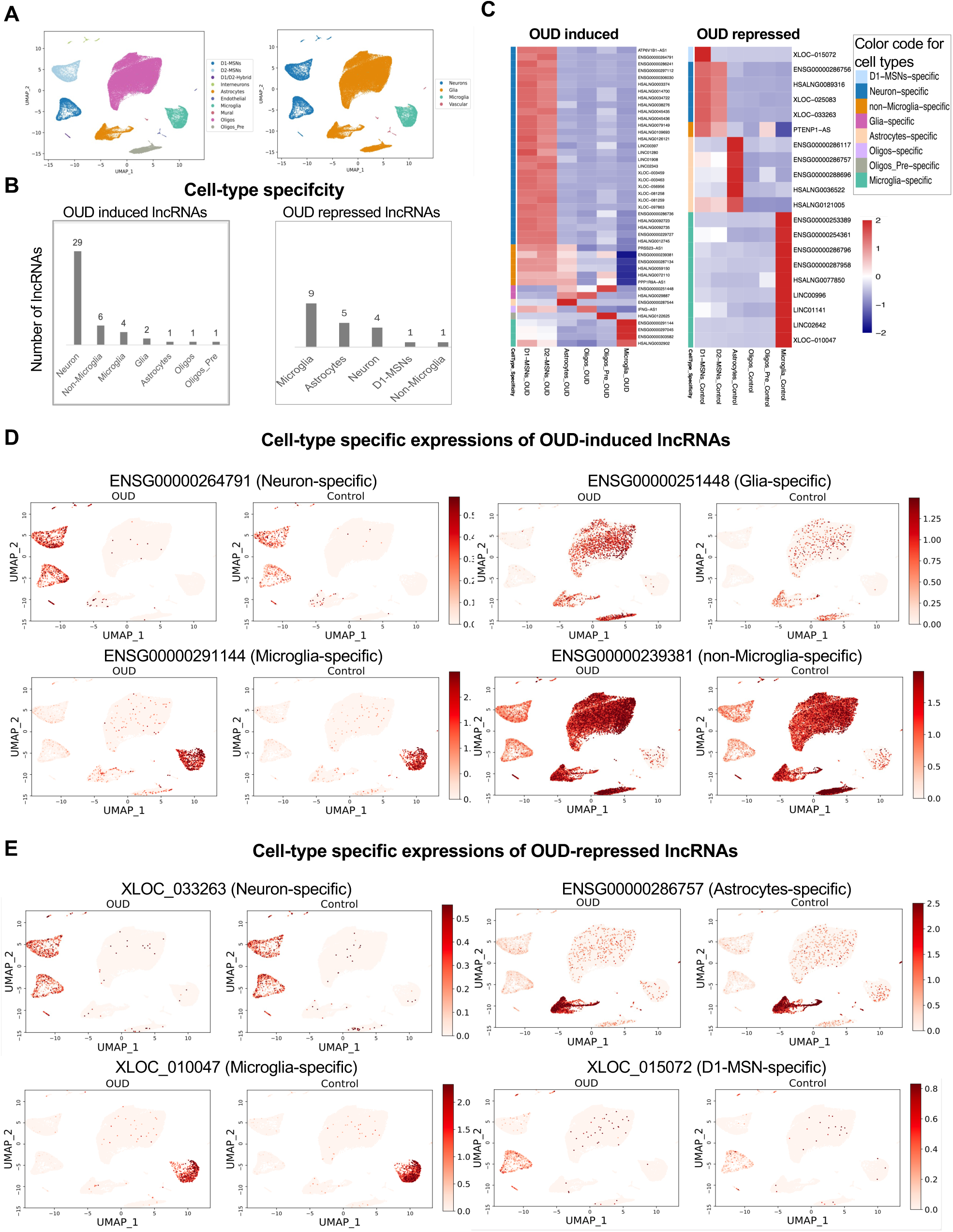
OUD-associated lncRNAs exhibit cell type-specific expression patterns in the striatum. (A) UMAP visualization of striatal cell populations. Cell-type annotations were adopted from our prior single-nucleus RNA-seq study (Phan et al., *Nature Communications* 2024). Major cell classes are displayed either individually (left) or grouped into broad categories (right), including neurons (D1- and D2-MSNs, and inter-neurons), glial cells (astrocytes, oligodendrocyte lineage cells), vascular cells (mural and endothelial), and microglia. (B) Number of OUD-dysregulated lncRNAs in NAc, a major component of ventral striatum, showing cell type-specific expression across major stratal cell populations. Cell type-specificity was determined using snRNA-seq data from dorsal striatum, which shares highly similar cellular composition with ventral striatum. (C) Heatmap showing cell-type coverages of these cell type–specific lncRNAs across major striatal cell types. OUD samples were used to depict cell coverage of OUD-induced lncRNAs (left), whereas non-OUD samples were used to depict cell coverage of OUD-repressed lncRNAs (right). Rows represent cell type–specific lncRNAs, and columns represent the six major cell types in the striatum. (D) UMAP visualization of representative OUD-induced lncRNAs exhibiting cell type–specific expression patterns, illustrating selective activation within specific neuronal or microglia or glial populations. (E) UMAP visualization of representative OUD-repressed lncRNAs exhibiting cell type–specific expression, highlighting expression within defined striatal cell types.

One notable exception was HSALNG0121005, an OUD-repressed lncRNA. HSALNG0121005 exhibited an astrocyte-enriched expression in unaffected controls, but appeared relatively more neuron-enriched in OUD, suggesting a shift in cell type–specific expression upon opioid addition.

We further illustrate representative examples using UMAP plots. Among OUD-induced lncRNAs, ENSG00000264791 was primarily neuronal (including both D1- and D2-MSN) in the striatum **(Figure 5D, top left)**, whereas ENSG00000251448 was enriched in glia lineages (astrocytes, oligodendrocyte precursors, and oligodendrocytes), with stronger signal in OUD **(Figure 5D, top right)**. Microglia-specific expression is exemplified by ENSG00000291144 (**Figure 5D, bottom left)**. Conversely, some OUD-induced lncRNAs showed minimal expression in microglia yet broad expression across other cell types, such as ENSG00000239381 **(Figure 5D, bottom right** OUD-repressed lncRNAs also exhibited significant microglia-, astrocyte-, and neuron-enriched patterns (**Figure 5C, right**), with representative examples shown in **Figure 5E**. Notably, a D1-MSNs–specific expression pattern was observed among OUD-repressed lncRNAs (**Figure 5E, bottom right**), whereas none of the OUD-induced lncRNAs showed strong D1- or D2-MSNs specific expressions **(Figure 5E, fourth panel)**.

Together, these analyses support that a subset of OUD-associated lncRNA was dysregulated and organized in a highly cell type–resolved manner within striatal circuitry, implicating both neuronal and immune/glial compartments.

## DISCUSSION

### The noncoding genome is extensively remodeled in opioid use disorder

In this study, we provided a comprehensive characterization of long noncoding RNAs (lncRNAs) in human cortico–striatal circuits and demonstrate that the noncoding transcriptome is extensively remodeled in opioid use disorder (OUD). By systematically identifying annotated and previously unannotated lncRNAs in postmortem nucleus accumbens (NAc) and dorsolateral prefrontal cortex (DLPFC), we revealed widespread OUD-associated lncRNA dysregulation at multiple levels: steady-state expression, co-expression network organization, circadian rhythmicity, and cell type specificity. These findings extend prior transcriptomic studies of OUD, which have primarily focused on protein-coding genes, and establish lncRNAs as integral components of addiction-relevant molecular programs.

Approximately half of the lncRNA loci expressed in NAc and DLPFC were not represented in current annotations, underscoring the incomplete characterization of the human brain noncoding transcriptome. The high proportion of novel isoforms further suggests that transcript diversification is a prominent feature of cortico–striatal circuitry. Given the regional and cellular heterogeneity of these brain areas, it is likely that many of these lncRNAs are expressed in restricted neuronal or glial populations and have therefore remained undetected in annotation-dependent methods. Our findings emphasize that systematic lncRNA discovery is essential for a complete molecular understanding of addiction-related brain pathology.

### lncRNAs integrate neuroimmune and synaptic signaling programs in reward circuitry

In the NAc, we identified 375 differentially expressed lncRNAs in OUD, nearly half of which were novel. lncRNA-centered co-expression modules converged on pathways previously implicated in addiction neurobiology, including neuroimmune signaling, PI3K–AKT–mTOR cascades, GPCR downstream signaling, and focal adhesion processes. These pathways are central to synaptic remodeling, dopamine signaling modulation, and structural plasticity—key adaptations underlying compulsive drug seeking.

Importantly, the integration of lncRNAs within these modules suggests that they may contribute to organizing or stabilizing addiction-related transcriptional programs rather than merely reflecting downstream consequences of protein-coding gene changes. lncRNAs are known to scaffold chromatin modifiers, regulate transcription factor accessibility, and modulate RNA stability^19,31–35^. Thus, OUD-associated lncRNAs embedded within these modules may participate in fine-tuning inflammatory and synaptic responses in reward circuitry.

The enrichment of immune-related terms within NAc modules further supports accumulating evidence that neuroimmune activation contributes to opioid-induced plasticity. Whether lncRNAs function upstream of these immune processes or are themselves responsive to inflammatory signaling remains an important mechanistic question for future study.

### Executive circuitry exhibits distinct lncRNA-linked adaptations

In the DLPFC, we identified 102 dysregulated lncRNAs, forming modules enriched for phosphorylation-dependent signaling and intracellular receptor cascades. Notably, phosphorylation-centered processes emerged in both NAc and DLPFC, suggesting that kinase-regulated signaling is a shared molecular axis across reward and executive control circuits. Given that opioid receptor activation ultimately converges on intracellular signaling cascades that influence synaptic strength and plasticity, lncRNA association with these pathways points to potential roles in modulating circuit excitability and synaptic regulation.

However, DLPFC modules uniquely showed enrichment for cell cycle–related processes not observed in NAc. Because mature cortical neurons are post-mitotic, these signatures likely reflect glial or other non-neuronal adaptations in executive circuitry. Chronic opioid exposure may therefore engage distinct cellular responses in DLPFC compared to NAc, consistent with the differential functional roles of these regions in impulsivity, decision-making, and relapse vulnerability. These region-specific lncRNA networks suggest that reward and executive circuits undergo partially divergent noncoding regulatory remodeling in OUD.

### Circadian reorganization extends to the lncRNA transcriptome

Circadian disruption has emerged as a hallmark of substance use disorders, influencing reward sensitivity, mood regulation, and relapse risk. Here, we demonstrate that OUD disrupts rhythmicity of lncRNAs to a degree comparable to—and in some cases exceeding—that of mRNAs. Both gain- and loss-of-rhythmicity patterns were observed, indicating active reprogramming of oscillatory networks rather than uniform attenuation. In NAc, rhythmicity-altered lncRNAs were embedded within modules enriched for circadian clock components and phosphorylation pathways, linking temporal regulation with synaptic signaling cascades. In DLPFC, rhythmic modules converged on ion transmembrane transport, neuroactive ligand–receptor interactions, and retrograde endocannabinoid signaling—processes that directly modulate excitatory–inhibitory balance and neuromodulation.

The strong co-expression between rhythmic lncRNAs and neurotransmission-related genes such as GRIN2B and GABRA2 suggests that lncRNAs may contribute to the temporal coordination of excitatory and inhibitory signaling. Given that cortico–striatal communication depends on tightly regulated oscillatory dynamics, disruption of lncRNA rhythmicity may represent a systems-level mechanism linking molecular timing alterations to behavioral dysregulation in OUD.

### Cell type–specific lncRNA dysregulation highlights neuronal and glial contributions

A defining property of lncRNAs is their cell type specificity. By integrating single-nucleus transcriptomic data from striatum, we demonstrate that OUD-associated lncRNAs exhibit pronounced neuronal and glial enrichment patterns. The majority of OUD-induced lncRNAs were neuron-enriched, consistent with roles in synaptic remodeling and activity-dependent plasticity. In contrast, many OUD-repressed lncRNAs were enriched in microglia or astrocytes, suggesting altered neuroimmune and homeostatic regulation.

Subtype-specific patterns, including D1-medium spiny neuron–restricted repression, further indicate that lncRNA dysregulation may contribute to biased circuit remodeling within striatal pathways. Because D1- and D2-MSNs differentially regulate reinforcement and behavioral output, cell type–specific lncRNA regulation may shape functional imbalances in reward circuitry.

The observed shift in cell type–biased expression for specific lncRNAs suggests that OUD may alter not only expression magnitude but also cellular context of lncRNA activity. These findings reinforce the necessity of single-cell–resolved approaches to fully interpret noncoding transcriptome dynamics in neuropsychiatric disease.

### An integrated model of spatial, temporal, and cellular dysregulation

Taken together, our findings support a multi-dimensional model of lncRNA-mediated dysregulation in OUD. Spatially, lncRNA networks are differentially reorganized in reward (NAc) and executive control (DLPFC) circuits. Temporally, circadian rhythmicity of lncRNAs is reshaped, altering the timing of synaptic and signaling processes. Cellularly, dysregulation is partitioned across neuronal and glial populations, reflecting distinct contributions to synaptic plasticity and neuroimmune adaptation. These spatial, temporal, and cellular dimensions likely interact to destabilize cortico–striatal circuit function, contributing to compulsive drug seeking and impaired executive control.

### Limitations and future directions

This study leverages human postmortem tissue, providing direct disease relevance but capturing end-stage molecular states influenced by chronic exposure and potential acute drug effects. Functional validation of prioritized lncRNAs will be required to establish mechanistic roles in synaptic regulation and behavior. Future studies using perturbation models, temporal sampling, and region-specific single-cell transcriptomics will be essential to determine causal relationships. Additionally, investigation of conserved lncRNAs in rodent models may enable experimental interrogation of circuit-level mechanisms suggested by our network analyses.

## Conclusion

By integrating large-scale lncRNA discovery, expression profiling, network modeling, circadian analysis, and cell type–resolved transcriptomics, we demonstrate that long noncoding RNAs represent a critical and previously underexplored regulatory layer in opioid use disorder. Our findings redefine the addiction transcriptome to include dynamic noncoding regulatory programs and provide a framework for mechanistic investigation of lncRNA-mediated circuit dysfunction in OUD.

## METHODS

### Transcriptomic datasets

Poly(A)-selected RNA-seq data from 223 human nucleus accumbens (NAc) samples were obtained from GSE171936, and poly(A)-selected RNA-seq data from 30 dorsolateral prefrontal cortex (DLPFC) samples were obtained from GSE224683. These datasets were used for systematic identification and annotation of lncRNAs. For differential expression and rhythmicity analyses, ribo-minus RNA-seq data from NAc and DLPFC comprising 20 opioid use disorder (OUD) and 20 non-OUD individuals were retrieved from GSE174409. Single-nucleus RNA sequencing (snRNA-seq) data from dorsal striatum (GSE225158) were used for cell type–specific analyses. Two samples (GSM7040228 and GSM7040229) were excluded due to low sequencing quality^30^.

### Identification and annotation of lncRNAs

Novel and annotated lncRNAs were identified from poly(A)-selected RNA-seq data using *Flnc*^22^. All detected transcripts were merged into a unified annotation file (combined.gtf) using gffcompare tool^36^.

To classify transcripts, the merged annotation was first compared against GENCODE v47 (hg38)^25,26^ lncRNA annotations using gffcompare (-r). Transcripts assigned class codes other than “i”, “u”, or “x” were classified as GENCODE-annotated lncRNAs. Transcripts with codes “i”, “u”, or “x”, indicating no exon-level overlap with GENCODE annotations, were subsequently compared against LncBook v2.0 ^27^.Transcripts reassigned class codes other than “i”, “u”, or “x” were classified as LncBook-annotated lncRNAs, whereas the remaining transcripts were defined as novel lncRNAs.

For genes with multiple isoforms, a gene was classified as annotated if any isoform was annotated, with GENCODE annotation prioritized over LncBook annotation. Genes for which all isoforms were novel were classified as novel lncRNA genes.

### Genomic classification of lncRNAs

Genomic annotations of protein-coding genes from GENCODE v47 were compared with merged lncRNA transcripts using BEDTools^37^ and gffcompare^36^. lncRNA transcripts were classified into three genomic categories based on positional and strand relationships to protein-coding genes: (1) divergent, defined as transcripts whose transcription start site (TSS) was located within ±2 kb of the TSS of a protein-coding gene on the opposite strand; (2) antisense, defined as transcripts overlapping a protein-coding gene on the opposite strand by at least one base pair; and (3) intergenic, defined as all remaining transcripts. For genes with isoforms assigned to different categories, divergent classification was prioritized over antisense. Genes with exclusively intergenic isoforms were classified as intergenic.

### Genome browser visualization

Strand-specific paired-end RNA-seq reads were aligned to the human reference genome (hg38) using HISAT2^38^. Strand-resolved coverage tracks were generated using deepTools (bamCoverage function)^39^, with RPKM normalization and 10 bp bin size. Strand-specific BigWig files were generated using forward and reverse strand filtering. Genome browser tracks were visualized using pyGenomeTracks (v3.9) ^40^, incorporating GENCODE v47, LncBook v2.0, and the merged lncRNA annotation.

### Differential expression analysis

Differential gene expression between OUD and non-OUD samples (20 vs 20) was performed separately for NAc and DLPFC using a two-sided Wilcoxon rank-sum test stratified by sex and age (gene ∼ condition | sex + age). P values were adjusted using the Benjamini–Hochberg method. Genes with adjusted P < 0.05 and ≥1.5-fold increase in OUD were defined as OUD-induced, whereas genes with adjusted P < 0.05 and ≥1.5-fold decrease were defined as OUD-repressed.

### Co-expression network construction and module detection

To infer potential functions of differentially expressed (DE) or differentially rhythmic (DR) lncRNAs, lncRNA-centered co-expression networks were constructed separately for NAc and DLPFC. For each DE or DR lncRNA, Pearson correlation coefficients were calculated between the seed lncRNA and all other genes in the expression matrix. For each seed, the top correlated genes were selected to construct a local network, with network size capped at 100 genes and no more than five co-expressed lncRNAs included to prevent network dominance by noncoding transcripts.

Protein-coding genes within each network were mapped to Gene Ontology (GO) biological process pathways from the GSEA database^41^. Pathway match rates were normalized by pathway size. A network similarity matrix was then generated using Pearson correlation across pathway enrichment profiles. Networks were grouped into modules according to similarity. Module inclusion required that (1) addition of a new network did not reduce within-module correlation by more than 20%, and (2) total module size did not exceed 500 genes. All analyses were performed using in-house R scripts.

### Module characterization and enrichment analysis

Genes within each module were compared with curated addiction-related gene sets, primarily derived from published QTL studies, and with a circadian gene list^42^. Enrichment rates were calculated for each module, and modules with the highest overlap with addiction- or circadian-related genes were selected for visualization and downstream analysis. Representative networks were visualized using Cytoscape^43^. Pathway enrichment analysis of module protein-coding genes was performed using Metascape^44^.

### Differential rhythmicity analysis

Differential circadian rhythmicity between OUD and control samples was assessed using the DiffCircaPipeline R package^45^, applied separately to NAc and DLPFC datasets. Genes with FDR < 0.05 were defined as differentially rhythmic. lncRNAs with altered rhythmicity were subsequently used as seeds to construct co-expression networks using the same procedures described above. Module detection and enrichment analysis followed identical criteria.

### Single-nucleus RNA sequencing analysis

A custom STAR genome index was generated using hg38 and a combined GTF integrating GENCODE v47 protein-coding genes and merged lncRNA annotations. snRNA-seq reads were aligned using STARsolo (v2.7.10a)^46,47^ in droplet mode with 10x whitelist matching (one-mismatch correction). Gene expression matrices were generated using the GeneFull feature to capture pre-mRNA transcripts. Unique molecular identifiers were deduplicated with one-mismatch correction, and multimapping reads were assigned using expectation–maximization. Putative empty droplets were filtered using the EmptyDrops_CR method. Filtered UMI matrices were imported into Seurat v4^48^ and concatenated across samples for downstream analysis.

### Identification and visualization of cell type–specific lncRNAs

Cell type annotations followed our prior study^30^ and included D1-MSNs, D2-MSNs, Astrocytes, Oligos, Oligos_Pre, and Microglia. These were additionally grouped into three broader categories: Neurons (D1-and D2-MSNs), Glial cells (Astrocytes and oligodendrocyte lineage cells), and Microglia. Cell type– specific lncRNAs were identified using one-sided Wilcoxon rank-sum tests comparing each target cell type against all others. An lncRNA was defined as cell type–specific if it met all the following criteria: (1) adjusted P ≤ 0.01, (2) ≥5-fold higher mean expression in the target cell type, and (3) detectable expression in ≥5% of cells in the target cell type.

Heatmaps displaying the proportion of expressing cells per cell type were generated using the *pheatmap* package in R with row scaling. UMAP feature plots were generated using Python. To reduce the influence of extreme outliers, UMAP color-scale was designed for expression values capped at the 95th percentile of non-zero UMI counts.

## Authors’ Contributions

CF, RWL, and CZ conceived the study. ZL, CF, RWL and CZ designed the study. ZL performed lncRNA identification, classification, differential expression, and single nucleus RNA-seq analyses, with assistance from PZ. CF performed co-expression network, circadian rhythmicity and pathway analyses. ZL, CF, RWL and CZ interpreted the results. ZL, CF, and CZ drafted the manuscript, with input and revisions from PZ and RWL. RWL and CZ jointly supervised the study.

## Data Availability

The RNA-seq datasets analyzed in this study are publicly available from the original repositories described in the Methods section. No new primary sequencing data were generated for this study. Processed data files and derived analysis results supporting the findings of this work are available from the corresponding author upon reasonable request.

## ACKNOWLEDGEMENTS

We thank Madeline Kuppe-Fish from UMass Chan for her valuable discussion on single nucleus RNA-seq analysis.

## Declaration of generative AI and AI-assisted technologies in the writing process

During the preparation of this work, the authors used generative AI tools to correct grammar and spelling, and to improve the English language readability. After using this tool, the authors reviewed and edited the content as needed and take full responsibility for the content of the published article.

